# Hospital based contact tracing of COVID-19 patients and health care workers and risk stratification of exposed health care workers during the COVID-19 Pandemic in Eastern India

**DOI:** 10.1101/2020.11.01.20220475

**Authors:** Durgesh Prasad Sahoo, Arvind Kumar Singh, Dinesh Prasad Sahu, Somen Kumar Pradhan, Binod Kumar Patro, Gitanjali Batmanabane, Baijayantimala Mishra, Bijayini Behera, Ambarish Dash, G Susmita Dora, Anand L, Azhar S M, Jyolsna Nair, Sasmita Panigrahi, Akshaya R, Bimal Kumar Sahoo, Subhakanta Sahoo, Suchismita Sahoo

## Abstract

**Introduction:** Contact tracing and subsequent quarantining of Health Care Workers (HCWs) is essential to minimize further transmission of COVID-19 infection. In this study, we have reported the yield of Contact Tracing of COVID-19 Patients and HCWs and risk stratification of exposed HCWs.

**Methodology:** This is a secondary analysis of routine data collected for contact tracing from 19th March to 31st August 2020 at All India Institute of Medical Sciences, Bhubaneswar, Odisha, India. HCWs exposed to COVID-19 infections were categorized as per the risk stratification guidelines and the high-risk contacts were quarantined for 14 days and tested on 7^th^ day from last day of exposure. The low risk contacts were encouraged to closely monitor their symptoms while continuing to work.

**Results:** Out of 3411 HCWs exposed to COVID 19 patients (n=269) and HCWs (n=91), 890 (26.1%) were high risk contacts and 2521 (73.9%) were low risk contacts. The test positivity rate of high-risk contact was 3.82% and for low risk contact was 1.90%. Average number of high-risk contacts was significantly higher; for admitted patients (6.6) as compared to HCWs (4.0) and outpatients (0.2), p value = 0.009; for patients admitted in non-COVID areas (15.8) as compared to COVID areas (0.27), p value < 0.001; and when clustering of cases was present (14.3) as compared to isolated cases (8.2); p value < 0.001. Trend analysis (15 days block period) showed a significant decline in number of mean numbers of high-risk contacts during the study period.

**Conclusion:** Contact tracing and risk stratification was effective and helped in reducing the number of HCWs going for quarantine. There was also a decline in high-risk contacts during study period suggesting role of implementation of hospital based COVID related infection control strategies. This contact tracing and risk stratification approach designed in the current study can also be implemented in other healthcare settings.

## Introduction

With 44 million confirmed cases and over one million confirmed deaths affecting all the countries across the world, COVID-19 is currently the largest pandemics of the century [1]. In India, the number of cases are more than 8 million and deaths are beyond one lakh [2].

As of 21 April 2020, countries reported to World Health Organization (WHO) that over 90, 000 health care workers (HCWs) were infected with COVID-19. This number may be significantly higher because of underreporting [3]. COVID-19 Infection among HCWs not only possesses risk of infection to their family members thus contributing to community spread but also to other HCWs and patients. Thus, apart from stringent infection control practices to reduce the exposure to infection, contact tracing and subsequent quarantine of HCWs is essential to minimize further transmission. Consequently, isolation after COVID-19 infection and quarantine following exposure to a confirmed case of COVID-19 can adversely reduce the availability of human resources. To mitigate the shortage of staff in hospitals, WHO and Centre for Disease Control and Prevention (CDC) have given recommendations to stratify the risk following exposure into two categories viz. low risk exposure and high-risk exposure [4,5]. Ministry of Health and Family Welfare (MOHFW), Government of India has also adopted these guidelines [6]. In our hospital which caters to both COVID-19 patients and other patients, we adopted the contact tracing and risk stratification approach based on these guidelines to categorize exposed HCWs into high and low-risk contacts. In this paper, we have reported the yield of hospital-based contact tracing of patients and HCWs tested positive for COVID-19 and risk stratification of exposed HCWs in the hospital a statutory body under the aegis of MOHFW, Government of India.

## Materials and methods

### Study design

This is a secondary analysis of routine data collected from 19^th^ March to 31^st^ August 2020 during the process of contact tracing and laboratory data related to COVID-19 at the study site.

### Study site

The study was conducted at All India Institute of Medical Sciences (AIIMS), Bhubaneswar, which is a 960 bedded tertiary care teaching hospital located in Bhubaneswar, the capital city of Odisha, an Eastern state of India.

### COVID-19 related clinical services at study site

Patients reporting to the hospital were screened for COVID-19 as per the screening algorithm depicted in Fig 1. COVID-19 screening with Reverse Transcription-Polymerase Chain Reaction (RT-PCR) test of all the newly admitted patients, irrespective of presence of symptoms, started from 15^th^ June 2020. On 19^th^ March 2020, the first patient (second case of Odisha) with COVID-19 was admitted in our hospital. Patients presenting with symptoms consistent with COVID-19 and confirmed cases of COVID-19 were admitted in separate dedicated wards called “COVID-19 ward or COVID area”. From July 10^th^ onwards, routine outpatient consultation was discontinued due to sudden surge in COVID-19 cases in community and hospital. Hospital admission was restricted to only COVID-19 patients and patients requiring emergency or essential intervention.

**Fig. 1:**
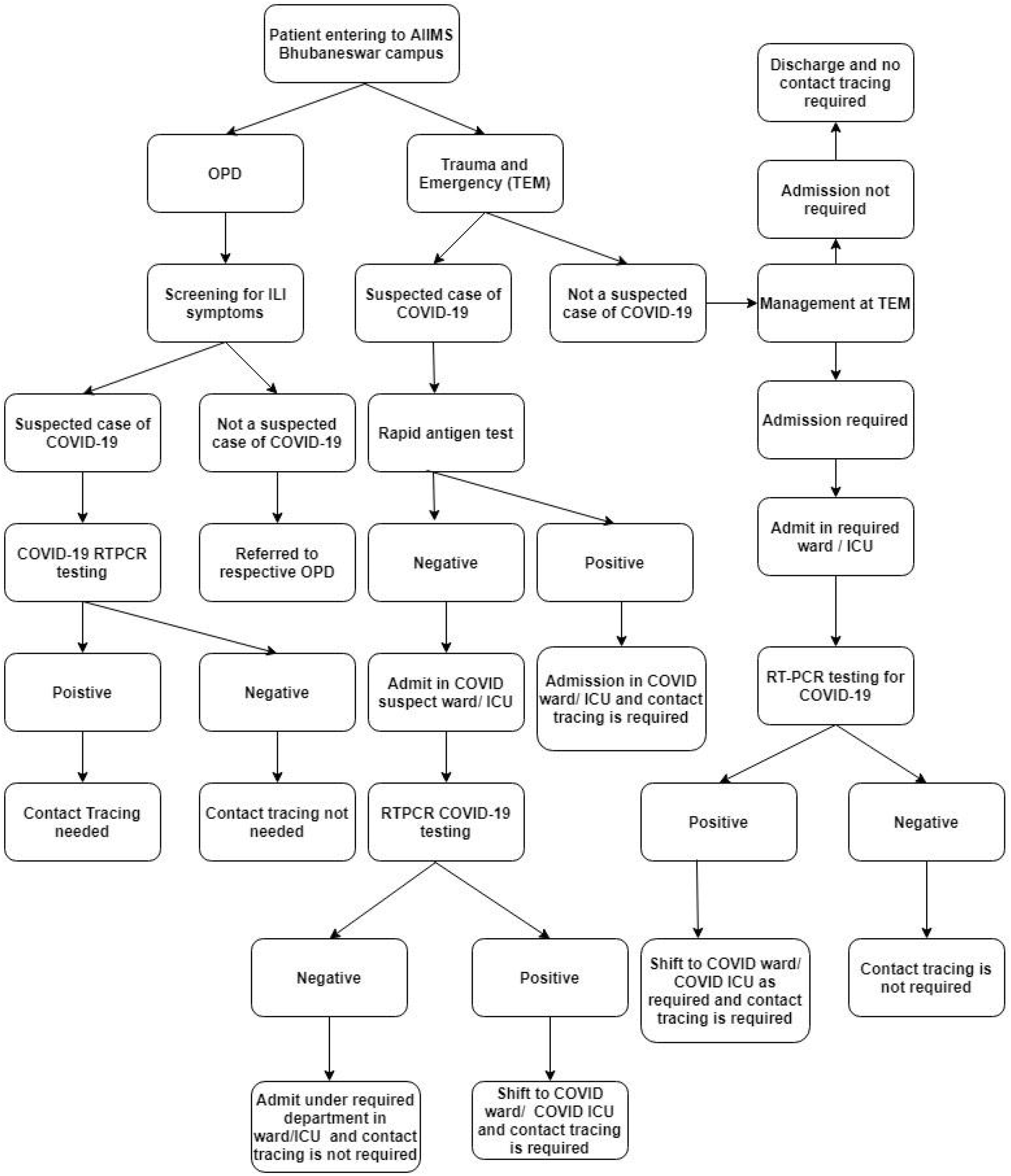
Algorithm for COVID-19 Testing strategy of Patient reporting to the hospital Study Procedures.

### COVID-19 related prevention interventions at study site

Various training programs were conducted to train all cadre of HCWs regarding proper use of Personal Protective Equipment (PPE), hand hygiene measures and other infection control practices. Use of type of PPE in different clinical areas and hospital premises was guided by MOHFW protocol and upgraded or modified based on feedback received from contact tracing or infection control team. Advisories were issued to all HCWs at periodic intervals for PPE compliance and infection control measures We also introduced various behavioral and regulatory interventions to promote COVID appropriate behaviors, for example monetary penalty for not using mask in hospital and residential campuses.

As per the testing strategy outlined in Fig 2, testing for SARS-CoV-2 using RT-PCR method was performed at COVID-19 RTPCR testing laboratory of institute, an approved laboratory by Indian Council of Medical Research (ICMR), Government of India. Contact tracing of laboratory confirmed COVID-19 patient, was initiated by a team dedicated for contact tracing immediately after intimation from diagnostic laboratory. Initially, contact tracing was done by physically visiting the clinical areas, personally interviewing the HCWs involved in patient care, reviewing medical records of patients, duty rosters and viewing CCTV footage. However, the strategy was modified with increase in number of cases to a passive mechanism of contact tracing. In later phase (from July 15 onwards), Contact Tracing Team (CTT) intimated the concerned departments to provide list of all HCWs who had possibly come in contact with confirmed COVID-19 cases in a prescribed format. Upon obtaining the list of exposed HCWs, the CTT contacted each HCW telephonically to elicit history related to duration and type of exposure, procedures performed on the patient, use of personal protective equipment during exposure etc. In case of contact tracing related to HCW being a confirmed case of COVID-19, history related to interactions during duty break hours, during meals, and other places where HCW is likely to be less cautious regarding mask usage was probed while contact tracing. Contacts in last 14 days from the date of positive report was considered for contact tracing. Risk categorization (low risk exposure and high-risk exposure) as per the criteria adopted using WHO, CDC and MOHFW guidelines as given in Box 1. A 14 days home quarantine and COVID-19 testing on 7^th^ day of last exposure was recommended for HCWs having high risk exposure whereas HCWs having low risk exposure were recommended to continue the work. The quarantine period was considered as fully paid on-duty period. Both the risk categories were required to monitor for symptoms and report for COVID-19 testing on appearance of symptoms consistent with COVID-19. In absence of symptoms, routine testing was not recommended for low risk contacts. However, few HCWs having low risk exposure were also tested based on their request. We collated data related to contact tracing and risk categorization in excel spreadsheet and follow up of HCWs was done to enquire about symptoms and test results. CTT regularly updated hospital authorities about their findings related to breach in infection control practices and areas of high-risk contacts and suggested specific recommendations. Ethical approval to conduct this study was obtained from Institutional Ethics Committee of AIIMS, Bhubaneswar. (Reference number: T/IM-NF/CMFM/20/76)

**Fig. 2:**
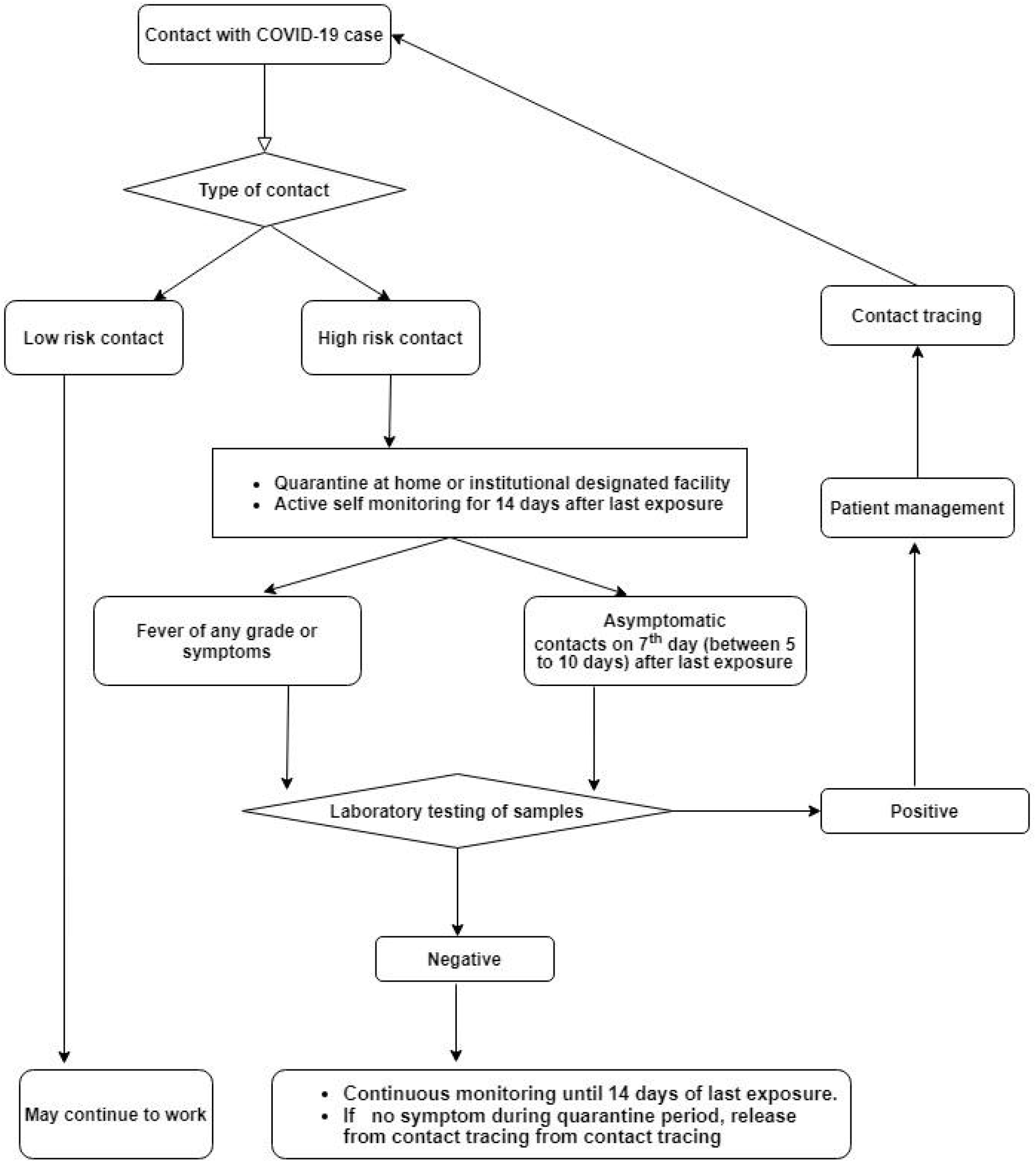
Testing strategy for SARS-CoV-2 after contact tracing Statistical Analysis.

#### Box 1

**Risk categorization (low risk exposure and high-risk exposure) as per the criteria adopted using WHO, CDC and MOHFW guidelines**

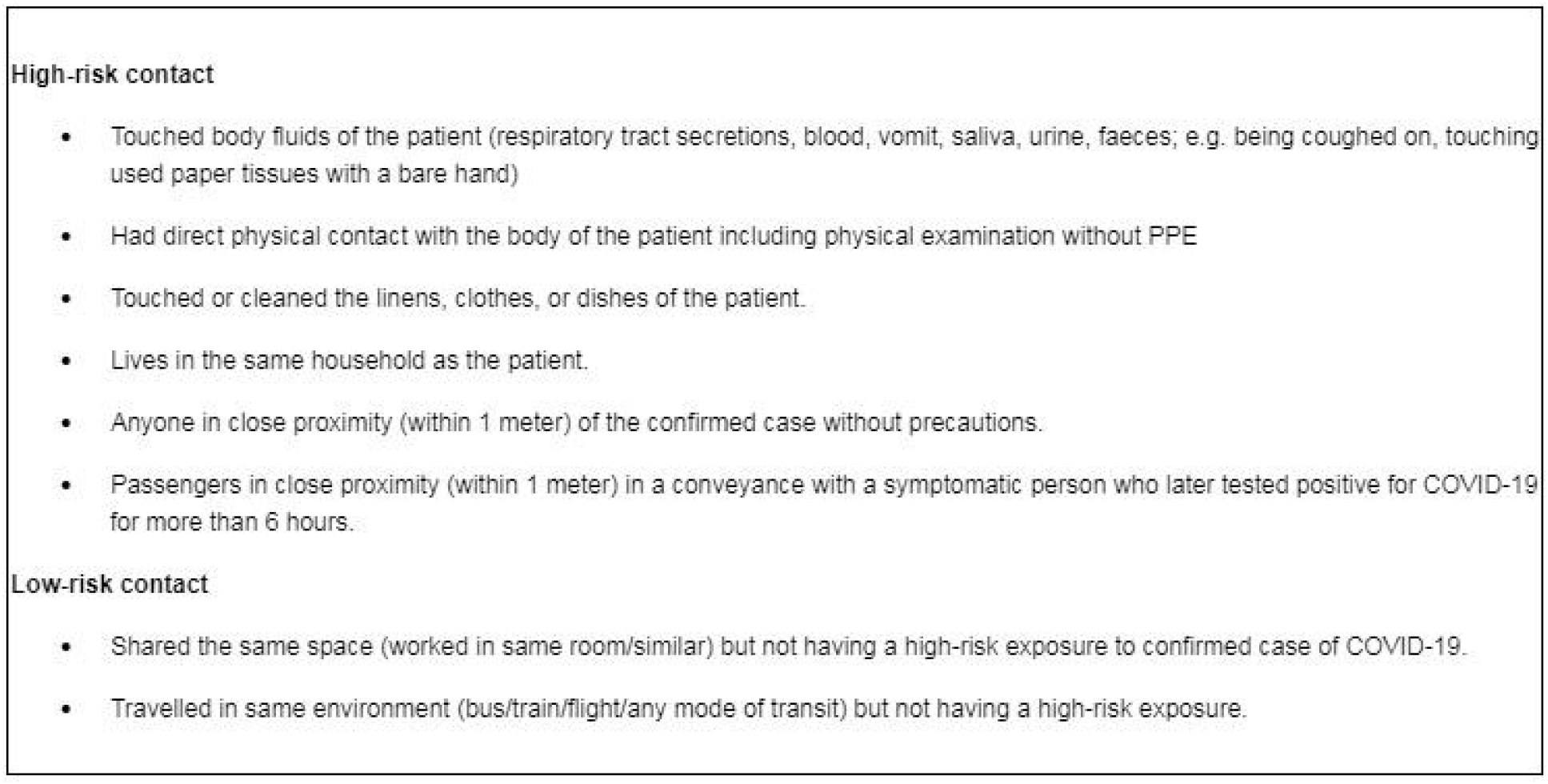

Statistical analysis was done using Microsoft excel 2013 version and Statistical Package for Social Science’s version 22.0 (SPSS 22.0). Descriptive statistics was presented as mean (± standard deviation) and percentage (± 95% confidence interval). Patients with COVID-19 were categorized into inpatient, outpatients and HCW tested positive for COVID-19. Inpatients were further categorized based on area where they were admitted i.e. COVID area or non-COVID areas. Clustering of cases was defined as diagnosis of more than two patients with COVID-19 on a single day from a non-COVID area. When a cluster was present, the total number of high risk and low risks were considered for each patient for comparison between groups and for descriptive analysis the overall high risk and low risk cases were considered. For example, if there was a cluster of 5 cases with 10 high risk contacts, then for comparison analysis 50 high risk contacts were considered while for descriptive analysis the count remains as 10. Mean number (± standard deviation) of high-risk contacts and low-risk contacts were compared with respect to type of patient (i.e. admitted patient in COVID area, admitted patient in non-COVID area, outpatient, HCW) and clustering of cases, time period of 15 days from the date of admission of first patient.

## Results

This analysis included data related to 360 COVID-19 patients reported during the study period which included 240 (66.7%) admitted patients or Inpatient Department (IPD), 29 (8.1%) Outpatient Department (OPD) and 91 (25.3%) HCWs. Out of 269 IPD and OPD patients, 163 (60.6%) were admitted directly to COVID-19 areas, 97 (36.1%) were admitted in non-COVID areas and rest 9 (3.3%) had stayed in both COVID and non-COVID areas. (Table 1). Out of the 97 cases diagnosed in non-COVID areas, 56 presented in a cluster; with one cluster each of 12, 10, 4, 3 cases, two cluster with 6 cases, and three cluster with five cases each.

**Table 1:** Distribution of COVID-19 positive patients.

|  | Frequency | Percent |
| --- | --- | --- |
| **Type of patient (n = 360)** |  |  |
| Inpatient Department (IPD) | 240 | 66.7 |
| Out Patient Department (OPD) | 29 | 8 |
| Health Care Workers (HCWs) | 91 | 25.3 |
| **Area; excluding staff (n = 269)** |  |  |
| COVID area | 163 | 60.6 |
| Non-COVID area | 97 | 36.1 |
| Both | 9 | 3.3 |
| **Clustering of cases (n =56), number of patients in the cluster** |  |  |
| 12 | 1 | 21.4 |
| 10 | 1 | 17.9 |
| 6 | 2 | 21.4 |
| 5 | 3 | 26.8 |
| 4 | 1 | 7.1 |
| 3 | 1 | 5.4 |

Contact tracing team identified 3411 HCWs who were exposed to any COVID-19 patient in the hospital. After risk categorization, 26.1% (n=890) were identified as high-risk contact and 73.9% (n =2521) were low-risk contact. (Table 2) Within 14 days of exposure with COVID-19 patient, 34/890 high risk contacts (3.8%, 95% CI; 0.027, 0.052) and 48/2521 low risk contacts (1.9%, 95% CI; 0.014, 0.025) tested positive for SARS-CoV-2.

**Table 2:** Test positivity rate of COVID-19 infection among the contacts.

| **Risk category** | **Frequency** | **Positive test result**<br>**n (%; 95%CI)** |
| --- | --- | --- |
| **High risk** | 890 | 34 (3.82, 2.72 to 5.24) |
| **Low risk** | 2521 | 48 (1.90, 1.46 to 2.52) |
| **Total** | 3411 | 82 (2.40, 1.92 to 2.98) |

Highest number of high-risk contacts was for patients in a non-COVID area (mean ± S.D; 15.8 ± 18.3) followed by when patient was a HCW (mean ± S.D.; 4.0 ± 5.6). Mean high risk contacts per patient was less than 1 if the patient was admitted in COVID area or was provided services on an outpatient basis. Difference between mean number of high-risk contacts in different groups was statistically significant (p<0.001) When the cases presented as clusters (i.e. more than 2 COVID-19 cases in one clinical area on a single day), mean number of high-risk contacts was 14.3 (± 19.4) whereas in case of isolated case it was 2.6 (± 5.9). The difference was statistically significant (p<0.001). (Table 3)

**Table 3:** Comparison of average high risk and low risk contact with respect to type of index case.

|  |  |  | Number of cases | Mean number of contacts | Std. Deviation | Unpaired T test/ Anova test | P value |
| --- | --- | --- | --- | --- | --- | --- | --- |
| **Type of patient** | High risk | IPD | 240 | 6.61 | 13.895 | 4.741<sup>#</sup> | 0.009 |
|  |  | OPD | 29 | 0.22 | 0.698 |  |  |
|  |  | Staff | 91 | 4.02 | 5.653 |  |  |
|  | Low risk | IPD | 240 | 10.81 | 11.754 | 8.527<sup>#</sup> | 0.002 |
|  |  | OPD | 29 | 3.07 | 2.541 |  |  |
|  |  | Staff | 91 | 8.12 | 6.789 |  |  |
| **Area** | High risk | COVID area | 163 | 0.27 | 1.207 | -10.853* | 0.0001 |
|  |  | Non-COVID area | 97 | 15.84 | 18.268 |  |  |
|  | Low risk | COVID area | 163 | 5.93 | 5.544 | -7.803* | 0.0001 |
|  |  | Non-COVID area | 97 | 16.19 | 15.188 |  |  |
| **Cluster** | High risk | Cluster present | 56 | 14.35 | 19.36 | 8.805* | 0.0001 |
|  |  | Absent | 304 | 2.65 | 5.904 |  |  |
|  | Low risk | Cluster present | 56 | 13.8 | 12.237 | 4.448* | 0.0001 |
|  |  | Absent | 304 | 8.21 | 9.433 |  |  |

<sup>#</sup> ANOVA test
\* Unpaired t test

Trend analysis (with 15 days block period) showed a significant decline in number of high-risk contacts. In case of HCW being the index case, mean number of high-risk contacts reduced from 12.7 during 1-15 June to 3.7 during 1-15 July and to 0.62 during 16-31 August (first case of HCW of hospital tested positive for COVID-19 on 2^nd^ June 2020). (Fig 3a and 3b) Similarly, for patient admitted in COVID area, number of high-risk contacts was 10.0 for the only patient admitted till 31^st^ March; which reduced to a mean of 1.0 during 1^st^ April to 15^th^ June and further to less than 0.1 after 15^th^ June. In non-COVID areas, mean number of high-risk contacts reduced from 31.5 (during 16-30 June) to 3.0 (during 16-31 July) and further to 0.7 (during 16-31 August). (Fig 3c and 3d)

**Fig. 3a:**
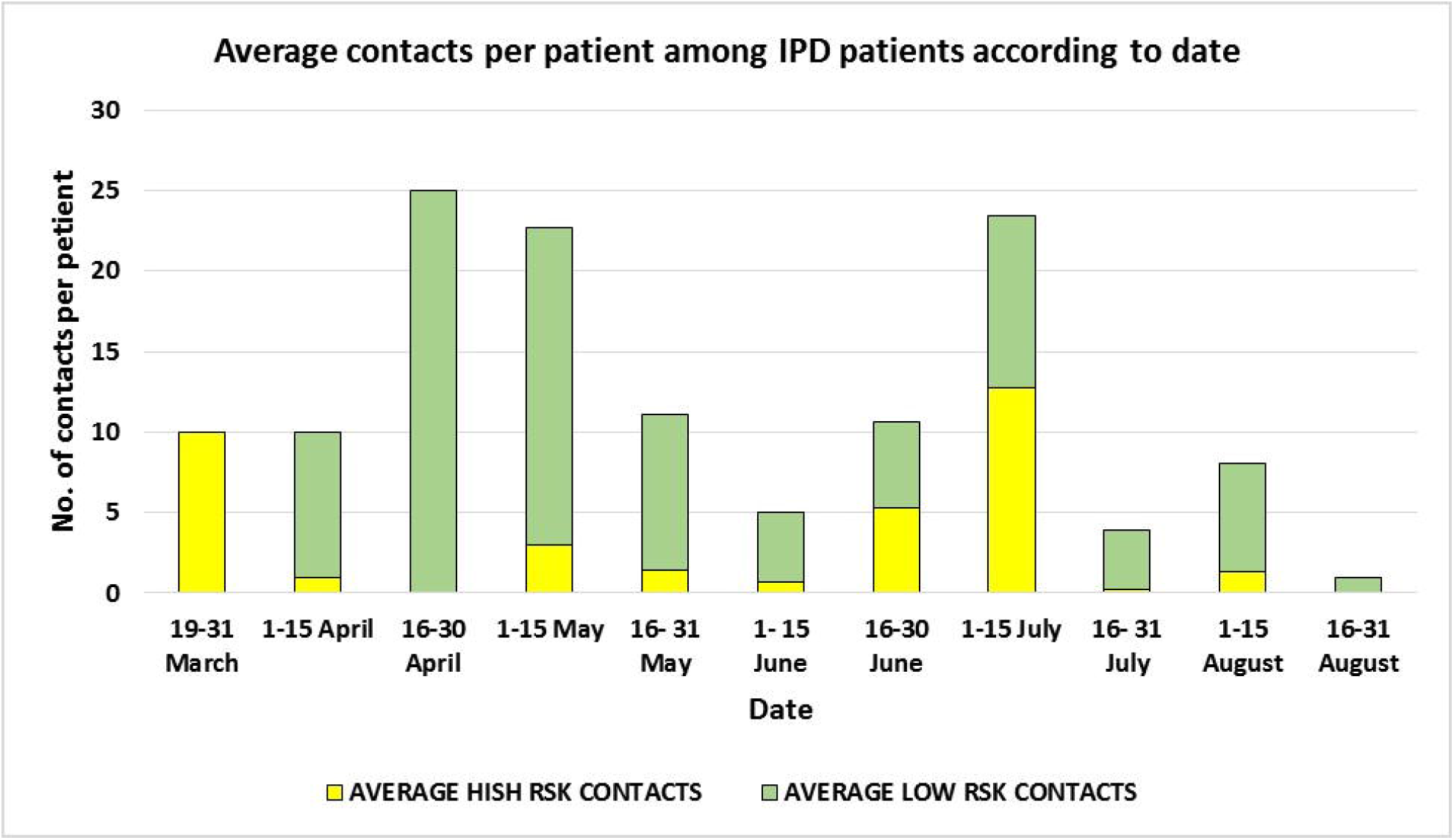
Time trend of average number of contact when COVID-19 patient was an admitted patient.

**Fig. 3b:**
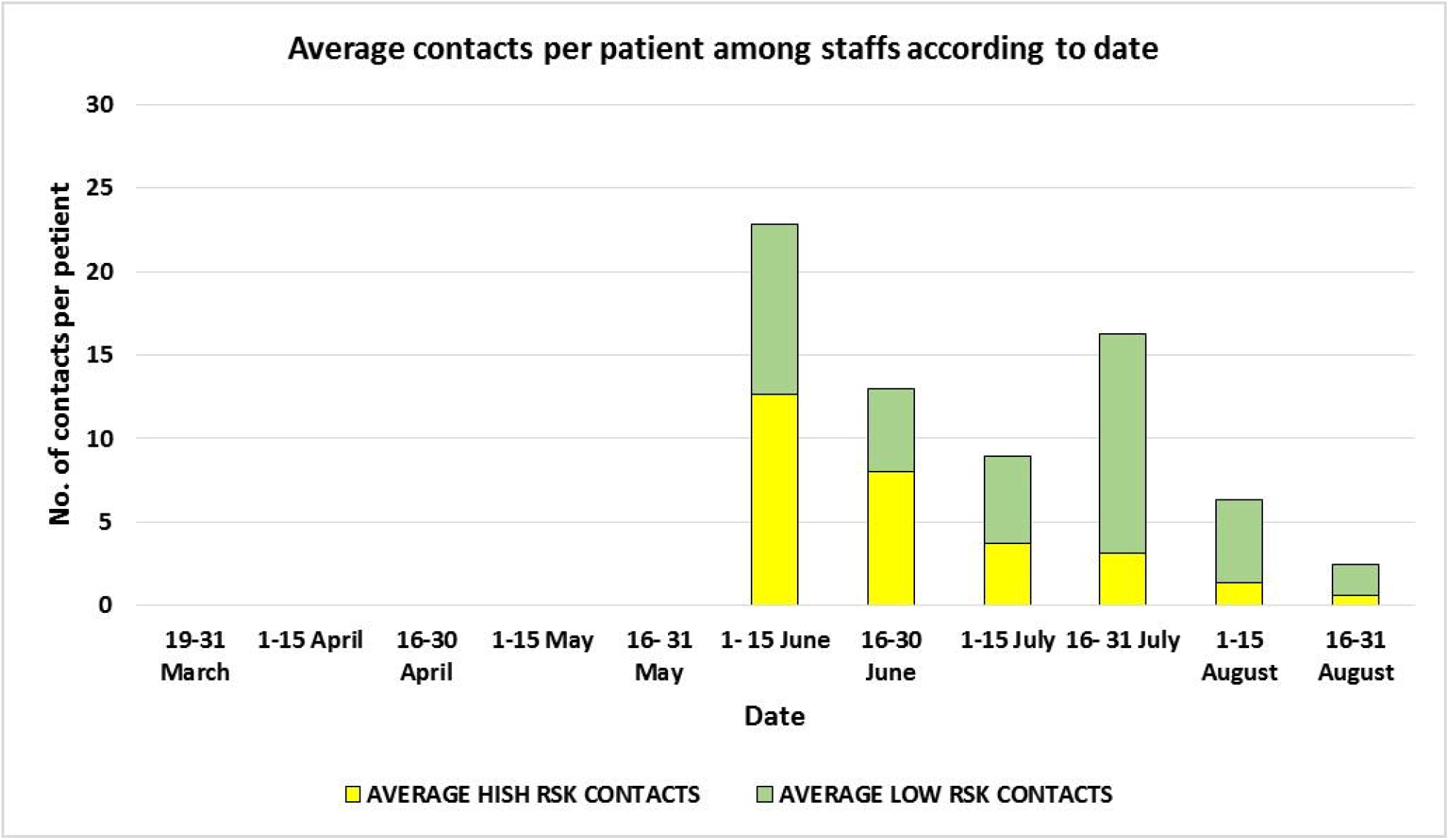
Time trend of average number of contact when COVID-19 patient was a Health Care Worker (HCW)

**Fig. 3c:**
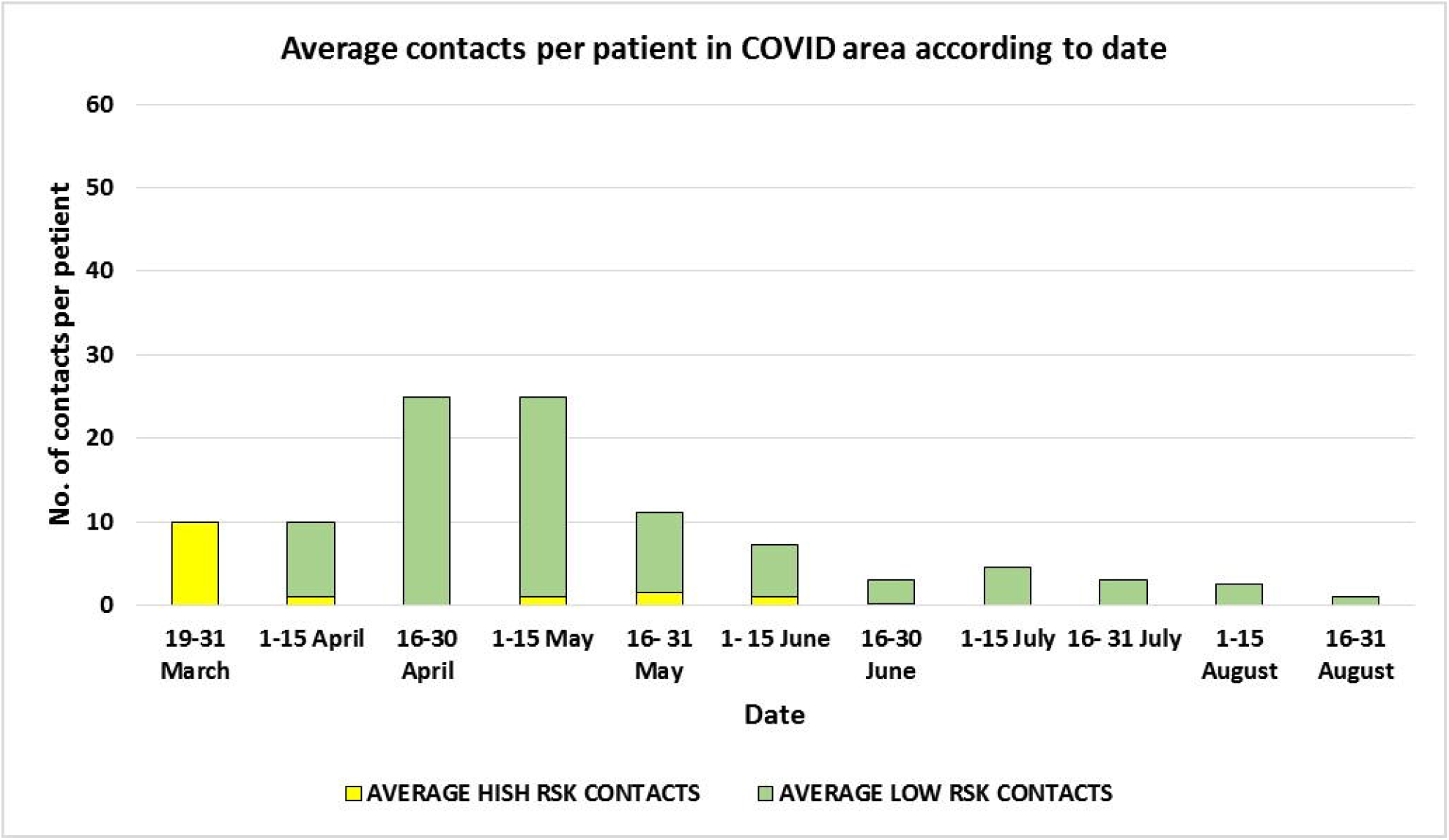
Time trend of average number of contact when COVID-19 patient was admitted in COVID area.

**Fig. 3d:**
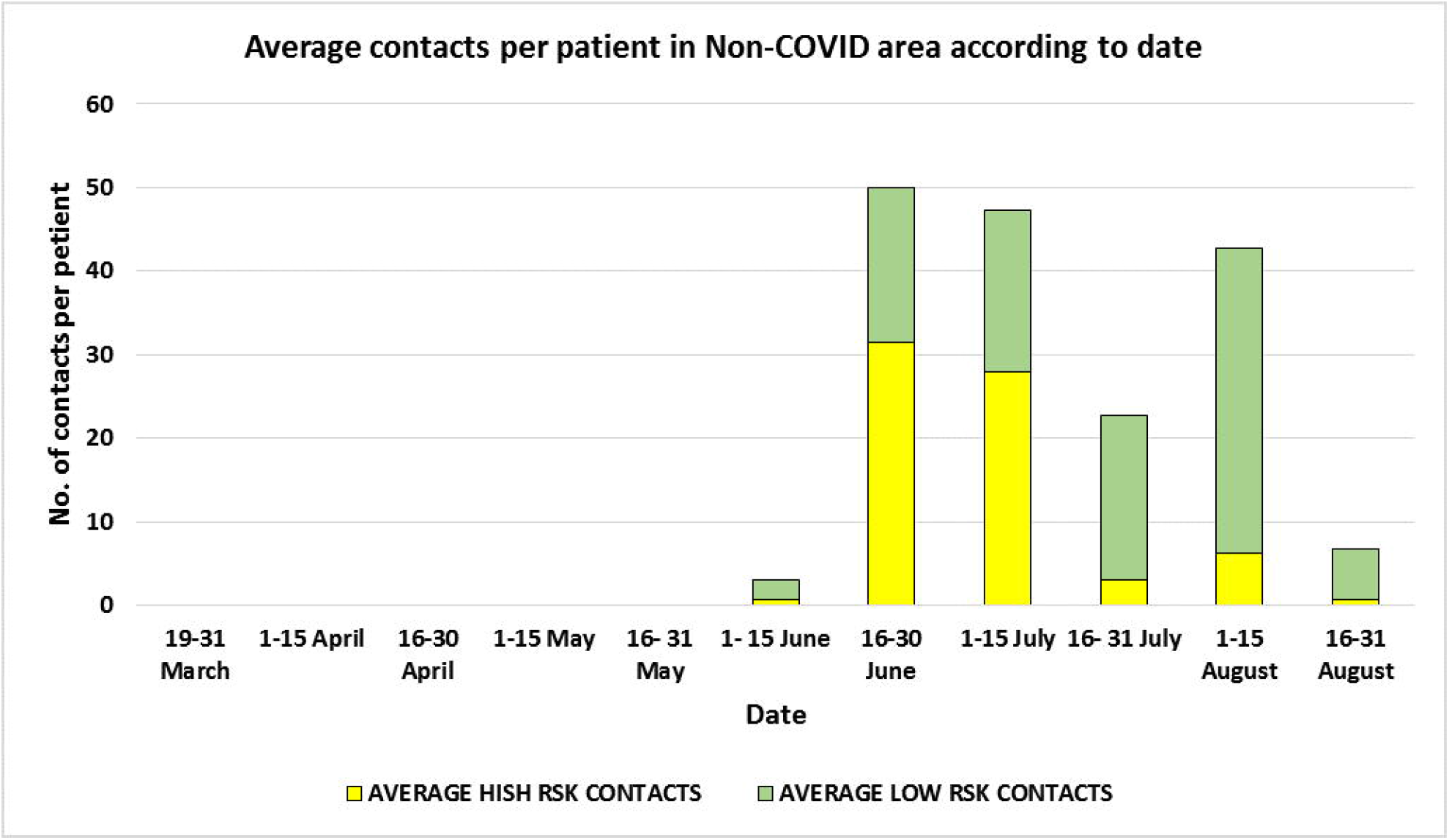
Time trend of average number of contact when COVID-19 patient was admitted in Non COVID area.

Interview with HCWs during contact tracing revealed that common causes for a high-risk exposure while providing clinical care was inadequate use of PPE and non-practicing of hand hygiene measures after direct contact with patient. Whereas, in case of a positive HCW, social interactions during meals and nursing stations during duty hours, hand over, travelling and staying together etc. contributed for majority of high-risk exposures

## Discussion

Our forward contact tracing of 360 patients and HCWs with COVID-19 encountered in the hospital during March-August 2020, revealed 3411 HCWs exposed to these patients. In our study, one fourth of exposed HCWs were stratified into high risk category and positivity rate for COVID-19 infection among them was 3.8% as compared to 1.9% among low risk contacts. We also observed a gradual decline in average number of high risk contact over the period of time. HCWs were more likely to be exposed when COVID-19 Infection was diagnosed among HCW and patient admitted in a non COVID area.

Few studies from India have also reported a similar proportion of high risk contact after risk stratification. In a study conducted in a hospital located in eastern part of India by Agarwal N et al where 25% of the exposed HCWs were high risk contacts [7]. However, this study had a small sample size. Another study conducted by Kaur R et al in a hospital located in northern part of India where only 14.5% of the exposed HCWs were categorized into high risk contacts. Our study reported a test positivity rate of 3.82% amongst high risk contacts while the study conducted by Kaur R et al observed higher test positivity rate of 7.1% [8]. In this study by Kaur et al, most of the COVID 19 patients (almost 85%) were HCW unlike our study where 25% of COVID-19 patients were HCW. Moreover, higher test positivity rate in this study could be due to more stringent criteria of stratifying into high risk category as proportion of high risk contact in this study was only 14% out of total exposed HCWs. Another study conducted by Blain H et al, observed high positivity rate amongst health care personnel (23.5%) but the study was conducted only for three COVID-19 positive cases [9]. In our study one index case had 10 high risk contacts on an average, while a study conducted by Vera C et al in Switzerland reported 21 high-risk contacts which was much higher as compared to our study [10]. This study from Switzerland was based on just one initially undiagnosed COVID-19 case.

We also observed a clear differential in the positivity rate among high-risk contacts and low-risk contacts (3.8% Vs 1.9%) which demonstrated the effectiveness of risk stratification strategy. Since, positivity rate among low risk contacts was also 1.9%, we ensured testing among low risk contacts, even if asymptomatic. Though, exposure to infection in low risk contact could also be from other sources as unlike high risk contacts, they were not quarantined and continued to work.

Number of high-risk contacts was highest when patient was admitted in non-COVID area (15.1) as compared to COVID area (0.3). This might be because, in COVID area HCWs were completely equipped with PPE while in non-COVID area they were with only surgical masks/N95 masks and or gloves as per the guidelines proposed by MOHFW, WHO and CDC and admitted patient was not a COVID-19 suspect. Since the admission was done on the basis of the history and signs and symptoms and after admission the testing was done to find out the RT-PCR status, during that period there was a high chance of exposure with the patients till the results of RT-PCR came.

The number of average high-risk contacts was higher (14.3) when there was a cluster of cases (2.6) as there was a chance that the whole department might be involved in patient care as the patients are more and hence there as compared to individual isolated cases. Similarly, the number of high-risk cases was more amongst admitted patients followed by HCWs and outpatients; as in IPD, the staffs perform their duties shift wise (three shifts in a day) as compared to OPD where there was only one shift. So, the chances of contamination were more. Also, a greater number of HCWs are engaged in clinical care activities in in-patient areas than in out-patient areas. The high-risk contacts of HCWs might be because of social mixing with the colleagues during the duty time and in residential areas. Thus, stringent infection control measures also need to be adopted in areas where patients are admitted which are not suspected of COVID-19.

Trend analysis showed a significant reduction in number of high-risk cases in all groups like COVID areas, Non-COVID areas, IPD, OPD and amongst HCWs, which reflects timely modification of infection control measures and strict implementation of PPE protocol and training of HCWs related to infection control which was implemented in hospital. Similar results were observed in a study conducted by Hidayat R et al in Indonesia in a COVID-19 referral hospital, where the secondary attack rate among HCWs declined with time from 20.1% to 3.7% [11]. However, at the same time there was decline in total exposed contacts (low-risk and high-risk combined) which might be indicative of either fatigue in contact tracing activity both in contact tracing team and the departments where COVID-19 patients were detected or human resource rationing (this included modification of duty rosters, less rotation of HCWs between different units) which was adopted as the pandemic progressed.

Contact tracing team based on enquiry from exposed HCWs also provided regular feedback to the hospital administration to augment infection control measures, identified areas of frequent breach in protocol, and suggested mechanism for reducing exposure to COVID-19. Apart from quarantine, regular feedback-based action might have helped in reducing exposure to COVID-19 infection in the hospital.

Contact tracing is a time and resource intensive exercise in both community settings as well as hospital settings. However, it is one of the most important tools for infectious disease prevention. In our hospital we used different methods described in literature for identifying the contacts exposed to COVID-19 patients like CCTV footage, duty rosters, and passive reporting by departments and telephonic enquiry. Contact tracing by data extraction from administrative and clinical databases such as electronic medical records (EMRs) or by CCTV footage (real-time locating system) have been reported previously [12,13]. Though, conventional contact tracing by continuous direct observation has been considered to be the gold standard method to accurately quantify contact time, however it requires intensive human resources and is not cost effective [14]. Self-reporting methods can be been used as alternatives to direct observation due to the lower intensity of their human resource demands. However, there is a chance of bias that compromise the accuracy of the data [15].

### Strengths

Since multiple strategies were employed like visiting the clinical area, personal interview of the HCWs, reviewing medical records, viewing CCTV footage etc., we expect that all the possible contacts were listed, tracked and categorized properly as it was performed by trained personnel and verified by experts. Thus, the quality of the data was expected to be satisfactory. Testing for COVID-19 was performed in an ICMR approved testing center by RTPCR which is considered as the gold standard. Continuous monitoring until 14 days of last exposure was done for all high-risk cases and testing was done on 7^th^ day.

### Limitations

Categorization of risk was based on the history by the contacts which may lead to high chances of social desirability bias. There was a chance of misinformation where the hospital staffs deliberately want to be in high risk categories so that a quarantine period of 14 days could be availed by them as paid on-duty leave. There was also a chance of misinformation due to wrong recall. Sometimes the HCWs fail to remember the patient and their PPE status during patient care. Low risk contacts were not tested as a routine unless symptomatic which could heave missed some cases as many of COVID-19 patients remain asymptomatic or paucisymptomatic.

### Conclusion

Contact tracing and risk stratification was effective and helped in reducing the number of HCWs going for quarantine. There was a decline in high-risk contacts during study period suggesting role of implementation of hospital based COVID related infection control strategies. Findings obtained during contact tracing might also be beneficial in mounting appropriate and strategic infection control measures. This contact tracing and risk stratification approach designed in the current study can also be implemented in other healthcare settings.

## Data Availability

Data

## Acknowledgement

We are grateful to the health care workers and the hospital administration for their co-operation during the contact tracing.

## Authors’ contributions

**Conceptualization:** Durgesh Prasad Sahoo, Arvind Kumar Singh, Binod Kumar Patro, Gitanjali Batmanabane

**Methodology:** Durgesh Prasad Sahoo, Arvind Kumar Singh, Binod Kumar Patro, Dinesh Prasad Sahu, Somen Kumar Pradhan, Gitanjali Batmanabane

**Data curation:** Durgesh Prasad Sahoo, Arvind Kumar Singh, Binod Kumar Patro, Dinesh Prasad Sahu, Somen Kumar Pradhan

**Investigation:** Durgesh Prasad Sahoo, Somen Kumar Pradhan, Dinesh Prasad Sahu, Baijayantimala Mishra, Bijayini Behera, Ambarish Dash, G Susmita Dora, Anand L, Azhar S M, Jyolsna Nair, Sasmita Panigrahi, Akshaya R, Bimal Kumar Sahoo, Subhakanta Sahoo, Suchismita Sahoo

**Formal analysis:** Durgesh Prasad Sahoo, Arvind Kumar Singh

**Supervision:** Arvind Kumar Singh, Binod Kumar Patro, Gitanjali Batmanabane

**Writing-original draft:** Durgesh Prasad Sahoo, Arvind Kumar Singh

**Writing-review & editing:** Binod Kumar Patro, Dinesh Prasad Sahu, Somen Kumar Pradhan, Gitanjali Batmanabane, Baijayantimala Mishra, Bijayini Behera, Ambarish Dash, G Susmita Dora, Anand L, Azhar S M, Jyolsna Nair, Sasmita Panigrahi, Akshaya R, Bimal Kumar Sahoo, Subhakanta Sahoo, Suchismita Sahoo

